# Publication Type Tagging using Transformer Models and Multi-Label Classification

**DOI:** 10.1101/2025.03.06.25323516

**Authors:** Joe D. Menke, Halil Kilicoglu, Neil R. Smalheiser

## Abstract

Indexing articles by their publication type and study design is essential for efficient search and filtering of the biomedical literature, but is understudied compared to indexing by MeSH topical terms. In this study, we leveraged the human-curated publication types and study designs in PubMed to generate a dataset of more than 1.2M articles (titles and abstracts) and used state-of-the-art Transformer-based models for automatic tagging of publication types and study designs. Specifically, we trained PubMedBERT-based models using a multi-label classification approach, and explored undersampling, feature verbalization, and contrastive learning to improve model performance. Our results show that PubMedBERT provides a strong baseline for publication type and study design indexing; undersampling, feature verbalization, and unsupervised constrastive loss have a positive impact on performance, whereas supervised contrastive learning degrades the performance. We obtained the best overall performance with 80% undersampling and feature verbalization (0.632 macro-F_1_, 0.969 macro-AUC). The model outperformed previous models (MultiTagger) across all metrics and the performance difference was statistically significant (*p <* 0.001). Despite its stronger performance, the model still has room for improvement and future work could explore features based on full-text as well as model interpretability. We make our data and code available at https://github.com/ScienceNLP-Lab/MultiTagger-v2/tree/main/AMIA.

## Introduction

Searching and filtering biomedical literature for relevant publications is a key step for learning about the latest advancements, conducting reviews, and synthesizing evidence. However, identifying relevant publications from the ever-growing literature can be a lengthy and cumbersome process^1–3^. To aid in this process, the U.S. National Library of Medicine (NLM) indexes MEDLINE articles by Medical Subject Headings (MeSH) and publication types (PT). For most of its existence, MEDLINE indexing has been performed manually by trained medical indexing experts^4^. In the last couple of decades, the Medical Text Indexer (MTI), developed at NLM, has increasingly automated the indexing process^5–8^, accelerating indexing from an average of 145 days down to a single day^9^. As of 2022, automatic indexing is applied to all journals indexed for MEDLINE, and human indexers review and curate the results^9^. Beyond the efforts at NLM, automated biomedical article indexing has received significant attention. BioASQ shared task competitions, which have taken place annually since 2013, focus on semantically indexing articles using MeSH terms with MTI commonly serving as a baseline^10–12^. Recent work on this task has explored deep learning architectures^13–15^.

MeSH headings and subheadings often correspond to the topical content of publications. Other groups have focused on implementing systems for commonly used publication types and study designs, which play an important role in searching and filtering for evidence synthesis. We note that while publication types sometimes align with a study’s experimental design, there can be differences. For example, an autobiography is a publication type but not a study design. On the other hand, random allocation is a study design, but it is not specific to any publication type. We will refer to all official publication types and study design-related MeSH terms as PTs, henceforth. PTs are distinct from many MeSH terms in that they indicate *how* the research was performed, rather than *what* the topic of the study is.

Due to the importance of randomized controlled trials (RCTs) in evidence synthesis, most of this research studied classification of RCT publications using machine learning^16,17^ or hybrid methods^18^. Cohen et al.^19^ have developed MultiTagger, a suite of 50 SVM classifiers that use features based on article title, abstract, and metadata to estimate the probability that an article has one or more of 50 PTs used in MEDLINE^20^. They computed PT cluster representations and used features about the journal of publication and other metadata through similarity comparisons with these clusters. MultiTagger achieved relatively high performance across a majority of PTs (44 labels with AUC greater than 0.90). In evaluation studies, RCT-Tagger^16^ and RCT-Tagger/MultiTagger combination have shown substantial work savings with minimal loss of included articles for systematic reviews^21,22^. Noting the inconsistencies in MeSH indexing, Cohen et al.^23^ also developed models for identifying human-related publications.

In this study, we hypothesize that state-of-the-art Transformer-based models^24^ combined with a multi-label task formulation could improve predictive power of PT tagging models, through enhanced text representations and the ability to leverage label correlations. To test this hypothesis, we construct a large dataset from PubMed abstracts and use PubMedBERT^25^ as the base encoder for PT tagging. We investigate whether verbalizing metadata features (e.g., journal name) and contrastive learning could further improve model performance, and compare our models to MultiTagger^19^.

### Related Work

Transformer-based models^24^, such as BERT^26^ and its variants trained on biomedical text^25,27^, have achieved state-of-the-art performance on many biomedical NLP tasks. In document classification, BioLinkBERT^27^ has shown state-of-the-art performance on the HoC (hallmarks of cancer) dataset^28^, commonly evaluated within the BLURB benchmark^25^. In work more related to our own, PubMed articles were classified by the experimental models used^29^. They annotated a dataset of 1,600 articles with 8 experimental models (e.g., *in vivo*, primary cells) and evaluated various biomedical BERT models with features derived from title, abstract, and MeSH terms, achieving the best performance with PubMedBERT^25^ (0.83 F_1_). They also trained models on abstract subsections; however, the model trained on the full abstracts outperformed the section-specific models.

BERT-based models leverage contextual information in text sequences to generate high-quality text embeddings. One strategy to embed non-textual metadata features in the vector space is to train separate embeddings for them and concatenate the results with the text embeddings. Another approach is to verbalize features before combining them with the text input. For example, Sainz et al.^30^ used verbalization to reformulate relation extraction as a classification task. With additional context regarding the task itself and the associated entities, they reported improvements in environments with less data.

Other techniques, focusing on model training, could also improve representations learned by the model. One such prominent approach is to use *contrastive learning*. First implemented for facial recognition^31^, this method has since been successfully applied to a variety of NLP tasks^32,33^. The general idea behind contrastive learning is that similar instances should have representations that are closer within the model embedding space, while dissimilar instances should be further apart. Contrastive learning has been studied in supervised^33^ and unsupervised^32^ settings. Unsupervised SimCSE method uses a variation of contrastive loss to achieve state-of-the-art sentence representations^32^. Lin et al.^33^ proposed several supervised contrastive loss functions specifically for the multi-label setting, which is more challenging than other contrastive learning due to the different levels of label overlap that can occur in this setting.

## Materials and Methods

### Dataset construction

We identified articles for inclusion using PubMed queries and downloaded them using the NCBI e-utilities API^34^. The base PubMed query restricted results to (1) English articles or articles with an English abstract, (2) articles published between 1987 and 2023, and (3) articles indexed by NLM involving a human in some way. Following MultiTagger^19^, for a subset of PTs (e.g., RCTs), we excluded articles with Editorial, Letter, Comment, Practice Guideline, or Review PTs. For some PTs (e.g., RCTs, Case Reports), we required the Humans MeSH term. All PT queries are available for download on GitHub. We used these human-curated terms as PT labels in this study. As an example, the query used for Clinical Trial PT is provided below.

> *((1987:2023[dp] AND (english[Language] OR english abstract[pt]) NOT indexingmethod automated) AND humans[MeSH Terms]) AND clinical trial[pt] NOT case-control studies[mh] NOT cohort studies[mh] NOT editorial[pt] NOT letter[pt] NOT comment[pt] NOT practice guideline[pt] NOT review[pt]*

As labels were available for more than 9 million PubMed articles, modified stratified sampling was used to restrict the dataset to a more reasonable size for training, while retaining enough samples for rare labels for effective model training. The stratified counts of all label combinations were calculated based on their proportion in the original set multiplied by the proposed size of the stratified dataset. Stratified counts below 1,000 were set to 1,000 or all available data if there were less than 1,000 instances of a label combination. Counts above 20,000 were capped at 20,000 to prevent any labels from dominating the dataset. The stratified dataset was supplemented with random negative articles (i.e., articles containing no positive labels) totaling 20% of the positive instances. Overall, these steps led to a dataset of 1,251,276 articles. These manuscripts were randomly stratified into 3 groups: training (70%, n = 875,892), validation (10%, n = 125,128), and testing (20%, n = 250,256).

### Feature extraction

We extracted titles, abstracts, and other metadata features from articles as model input. The extracted features and their frequency in the dataset are shown in Table 1. Not all features are present in every article. NCT identifiers and capitalized words were captured using regular expressions. Capitalized words are meant to capture abbreviations/acronyms (of RCT studies, for example). Features were verbalized in some experiments to better contextualize the data. When a feature was missing, the verbalized text was adjusted to reflect it (e.g., *This article’s publication date is unknown*.).

**Table 1:** The features used in the experiments, their frequency in the dataset, and their verbalization.

| Feature | % articles with the feature | Verbalization |
| --- | --- | --- |
| Title text | 99.9 | *This article’s title is ...* |
| Abstract text | 79.6 | No verbalization performed on this feature. |
| Journal name | 99.9 | *This article was published in ...* |
| Publication date | 99.6 | *This article was published in ...* |
| Keywords | 23.4 | *This article’s keywords are ...* |
| Number of references | 28.3 | *This article’s cited ... references* |
| Number of authors | 97.9 | *This article was written by ... authors* |
| Chemical list | 43.6 | *The chemicals mentioned in this article are ...* |
| Number of chemicals | 43.6 | *This article used ... chemicals* |
| NCT identifiers | 2.0 | *This article mentions the national clinical trial numbers ...* |
| Capitalized words | 58.7 | *This article uses the abbreviations ...* |

### NLP models

We used PubMedBERT^25^, a BERT-based model pre-trained from scratch on uncased, biomedical abstracts and full-text articles from PubMed Central, for our experiments. The overall experimental flow including a diagram of the model architecture is illustrated in Figure 1. Features extracted from PubMed articles were concatenated and fed as a single input to the PubMedBERT encoder. Inputs longer than the maximum sequence length (512 tokens) were truncated. The representation of the [CLS] token from this encoder’s last hidden layer was then regularized with dropout and fed into a linear layer with sigmoid activation for multi-label classification. Binary cross entropy loss (BCE) was used as the primary loss function.

**Figure 1:**
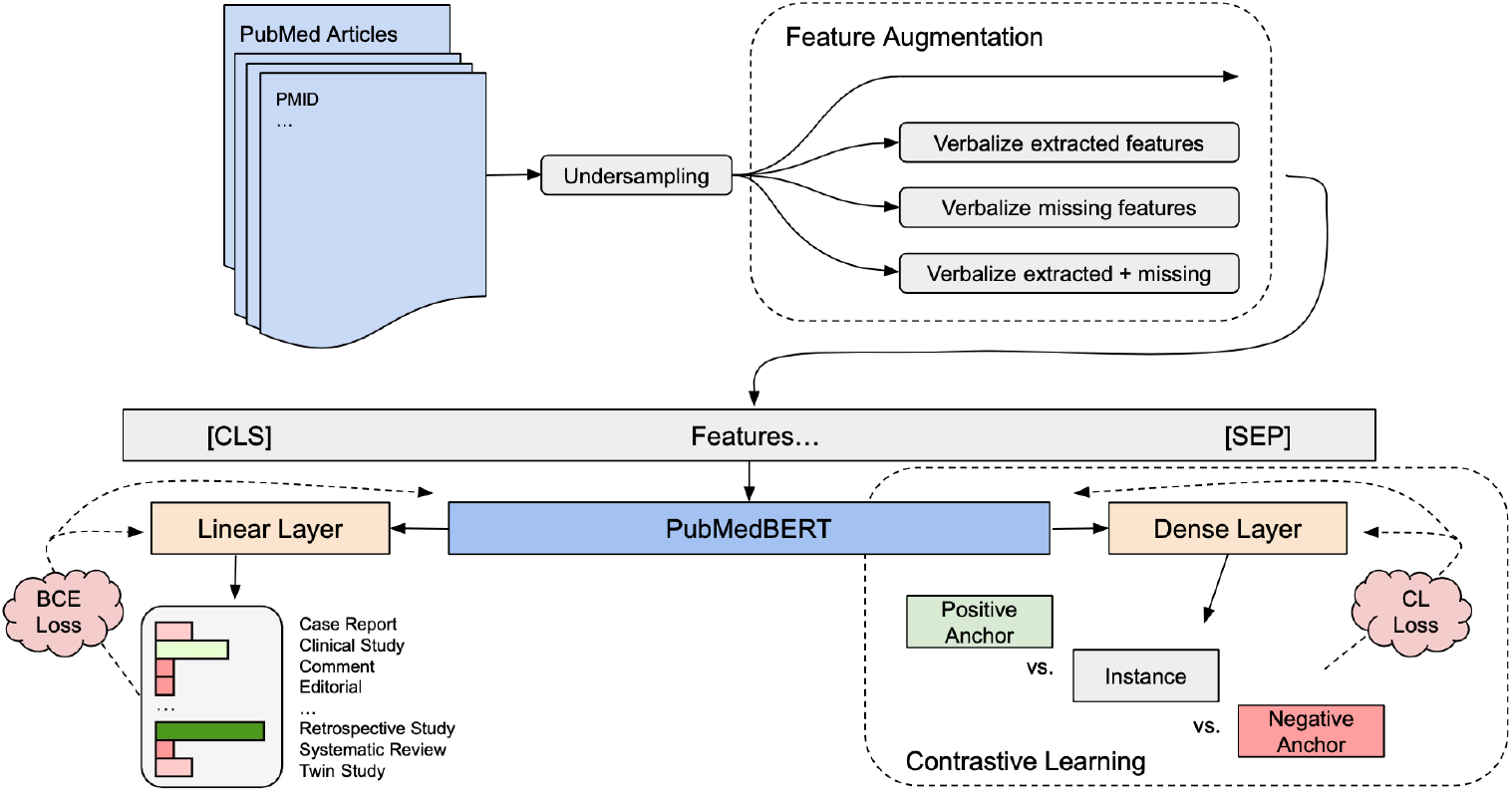
Flow diagram of experiments including data undersampling, feature augmentation, and contrastive learning. The dense layer used for contrastive learning experiments and the linear layer used for label predictions utilize the [CLS] token’s embedding from the last hidden state layer within PubMedBERT.

To reduce the computational resources required for training and to evaluate the effects of class imbalance, initial experiments tested model performance on subsets of the corpus. This involved setting, for each label combination, an *undersampling percentage* and the *minimum required number of instances*. Label combinations with more instances than the minimum were considered majority classes and were undersampled as necessary using the undersampling percentage (the proportion of removed instances). In experiments, we used undersampling percentages of 80%, 99%, and 99.9%, and set the minimum number of instances to 700 (for 80% and 99% undersampling) and 150 (99.9% under-sampling). In these experiments, all models were trained using verbalized data. In ablation studies, we also evaluated the effects of verbalizing present features only, verbalizing missing features, verbalizing all, and no verbalization.

We experimented with unsupervised and supervised contrastive loss. The unsupervised SimCSE method^32^ minimizes the difference between an instance’s representation with a noisy representation of itself, while maximizing the difference between itself and other samples. Our implementation of that loss function was as follows:

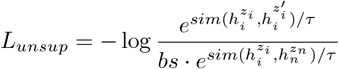

where *z*_*i*_ and *z*_*n*_ refer to the instance and its negative anchor, while 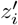 refers to the original instance (*z*_*i*_) with dropout used as noise; *h* refers to the hidden state representation, *sim* refers to cosine similarity, *τ* refers to the temperature hyperparameter, and *bs* refers to the batch size.

We also experimented with contrastive loss functions specific to the multi-label setting. We adapted strict contrastive loss (SCL) and Jaccard similarity contrastive loss (JSCL)^33^. SCL is defined as follows:

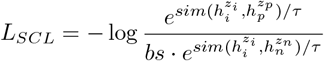

where *z*_*i*_, *z*_*p*_, and *z*_*n*_ refer to the instance, its positive anchor, and its negative anchor. While similar to SimCSE, for this loss, the positive anchor could be another instance rather than the instance itself. As some label combinations may be rare, our implementation incorporates noisy self-comparison (i.e., unsupervised SimCSE) to ensure there is at least one positive anchor from which to sample.

Finally, we also evaluated a combination of JSCL and SimCSE, wherein positive anchors were randomly selected from the set of instances with overlapping positive labels, as well as itself. The JSCL is defined here as follows, where Y_*i*_ and Y_*p*_ refer to the true positive labels associated with the instance and its positive anchor.

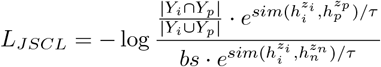

For contrastive loss experiments, we used a single positive anchor and a single negative anchor, rather than performing these loss calculations over mini-batches containing multiple negative anchors. The value associated with the negative anchor was then multiplied by the batch size rather than summing over the mini-batch^32,33^. This was done for SCL and JSCL as generating mini-batches with the correct number of positive and negative anchors would otherwise have been difficult, due to the label distribution in our dataset. For SimCSE, this was done to be more directly comparable with SCL and JSCL. Additionally, anchors were static; they were selected once at the start of training and reused over every epoch. This was done to reduce the computation required during training.

During these experiments, the model was trained jointly using BCE and contrastive loss (CL). The overall loss function is *L* = *α · L*_*CL*_ + (1 − *α*) *· L*_*BCE*_, where *α* is a hyperparameter controlling the preference between BCE and CL. For all CL experiments, we used an *α* of 0.4 to slightly prefer BCE over CL during training.

The following hyperparameters were used for model training and evaluation: batch size (32; for contrastive learning experiments, 16), learning rate (PubMedBERT layers: 1e-4; linear layers: 1e-2), linear learning rate scheduler, number of epochs (15), optimizer (AdamW^35^), dropout (0.1), and early stopping (when no improvement is observed on validation set macro-F_1_ over 2 epochs). All models were implemented using PyTorch (v.2.2.0) and PubMedBERT model weights downloaded from the HuggingFace transformers library^36^ (v.4.37.1). All experiments were conducted on a single Tesla v100 GPU with 32GB of memory.

### Model evaluation

Following MultiTagger^19^, we calculated optimal thresholds that maximize F_1_ score for each label on the validation set. Performance was then evaluated on the test set (n = 250,256) using these thresholds. We compared to MultiTagger on a subset of test set articles published in 2016 or later, where title and abstract were longer than 25 characters, using the 49 labels present in both models (n = 64,400 articles). We restricted by publication date to prevent data leakage, as MultiTagger was trained and calibrated using articles published between 1987 and 2015. We restricted by character length to provide a fair comparison as MultiTagger is unable to provide predictions on instances with fewer characters. We used standard evaluation metrics: precision, recall, and F_1_ score, as well as area under ROC curve (AUC). Each metric was micro- and macro-averaged; micro-averaging weighs each instance equally, while macro-averaging weighs each class equally. Macro-F_1_ was used as the main metric. We used one-sided, independent t-test to test whether there was a statistically significant difference in the model performances of MultiTagger and our models.

## Results

### Dataset statistics

The dataset consists of 1,251,276 articles. The average article within this dataset has 1.80 ± 1.29 PTs. The maximum number of PTs is 9 and the minimum is 0 (i.e., negative examples). The median number of PTs is 2, the lower quartile is 1, and the upper quartile is 3. Clinical Study is the most common PT in the dataset (n = 197k), with Prospective Studies (131k) and Follow-up Studies (116k) following closely behind. The three least frequent PTs are Scientific Integrity Review (n = 217), Veterinary Observational Study (n = 356), and Clinical Conference (n = 1,220). The label distribution for all 59 PTs predicted by our models is shown in Figure 2.

**Figure 2:**
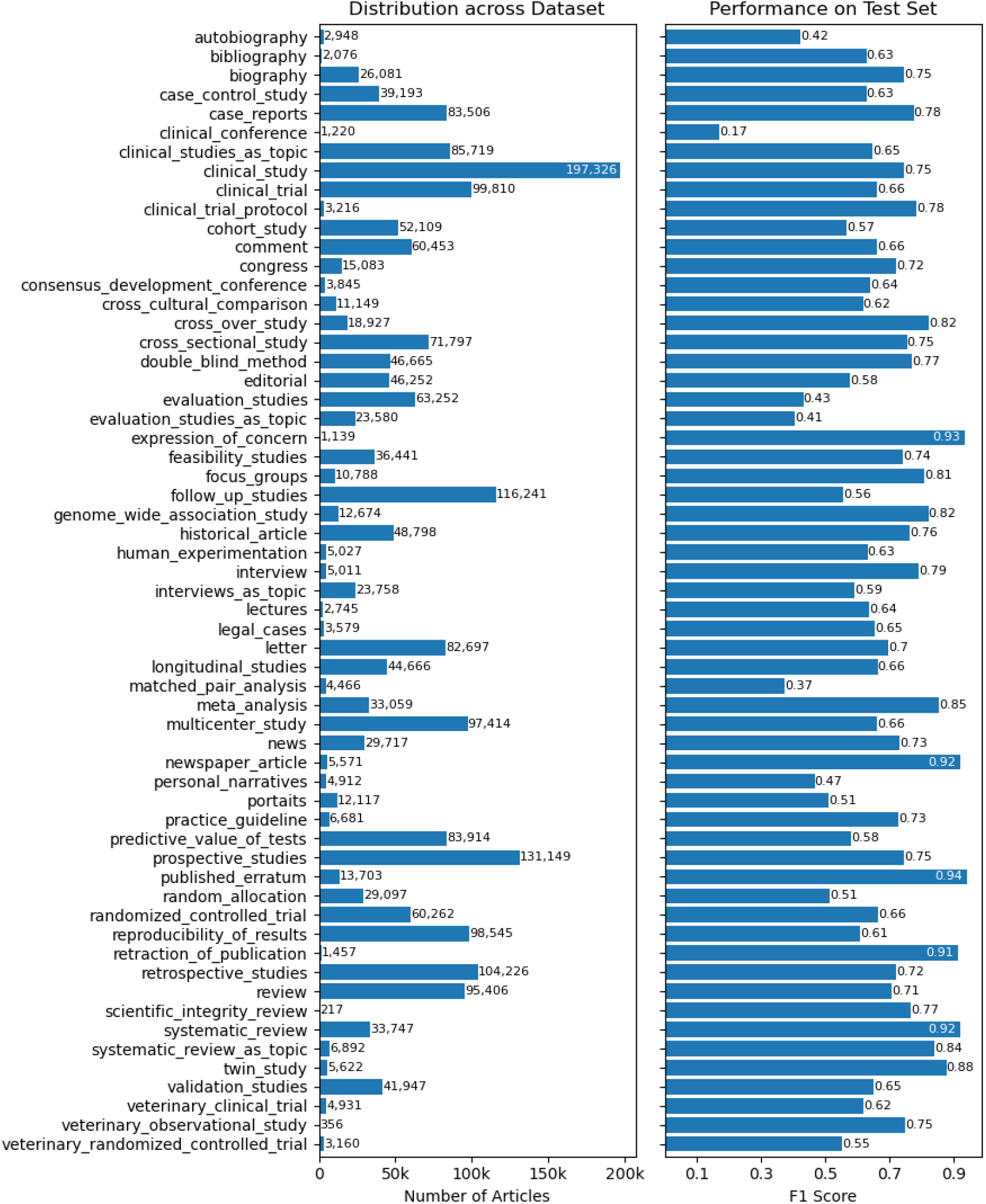
The sub-figure on the left shows the PT label distribution for all articles in the dataset. The right sub-figure shows the individual label performances using the best-performing model (80% undersampling and verbalized feature augmentation) on the test set.

### NLP models

The PubMedBERT models using feature verbalization yielded reasonable performance, reaching 0.681 macro-F_1_ score and 0.973 AUC with 80% undersampling. The effect of training set size on model performance is shown in Table 2. The differences between the 3 largest training sets (875k, 478k, and 413k), all using a minimum of 700 instances per PT, is relatively minor (macro-F_1_ range: 0.676-0.681). On the other hand, the smallest training set (189k) had a lower performance (macro-F_1_ = 0.661). This suggests that undersampling may be an effective way to reduce training size as long as the minimum number of instances for label combinations is set relatively high.

**Table 2:** Micro- and macro-averaged results on the test set for models using different training set sizes. Undersampling: undersampling percentage; Min.: minimum number of instances for label combinations. Best results are highlighted.

| Training Size | Undersampling | Min. | Precision |  | Recall |  | F<sub>1</sub> |  | AUC |  |
| --- | --- | --- | --- | --- | --- | --- | --- | --- | --- | --- |
|  |  |  | Micro | Macro | Micro | Macro | Micro | Macro | Micro | Macro |
| 875,892 | 0% | 700 | **0.667** | **0.695** | 0.680 | 0.671 | **0.673** | 0.676 | **0.980** | 0.972 |
| 477,793 | 80% | 700 | 0.666 | 0.692 | 0.679 | **0.682** | 0.672 | **0.681** | **0.980** | **0.973** |
| 413,113 | 99% | 700 | 0.656 | 0.688 | **0.683** | 0.681 | 0.669 | **0.681** | 0.979 | 0.972 |
| 189,279 | 99.9% | 150 | 0.634 | 0.666 | 0.670 | 0.665 | 0.651 | 0.661 | 0.977 | 0.970 |

The results of the comparison of our models with MultiTagger are shown in Table 3. We compared two models, PubMedBERT-189k as our baseline (no custom architecture or feature engineering) and PubMedBERT-Verbalized-478k as the best-performing model. Both models outperformed MultiTagger on the test subset, with PubMedBERT-Verbalized-478K achieving the highest macro-F_1_ score (0.632). The performance differences between our models and MultiTagger were statistically significant ((*p <* 0.001).

**Table 3:**
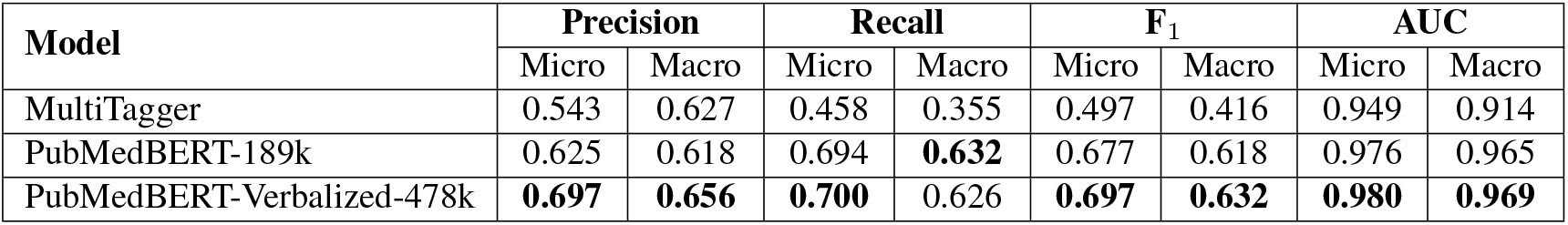
Comparison of our baseline and best-performing models with MultiTagger. The comparison used a subset of the test set (n = 64,400) to allow a fair comparison.

| Model | Precision |  | Recall |  | F<sub>1</sub> |  | AUC |  |
| --- | --- | --- | --- | --- | --- | --- | --- | --- |
|  | Micro | Macro | Micro | Macro | Micro | Macro | Micro | Macro |
| MultiTagger | 0.543 | 0.627 | 0.458 | 0.355 | 0.497 | 0.416 | 0.949 | 0.914 |
| PubMedBERT-189k | 0.625 | 0.618 | 0.694 | **0.632** | 0.677 | 0.618 | 0.976 | 0.965 |
| PubMedBERT-Verbalized-478k | **0.697** | **0.656** | **0.700** | 0.626 | **0.697** | **0.632** | **0.980** | **0.969** |

We also performed ablation studies for verbalization and contrastive loss functions. We limit these experiments to the smallest training set (189k) to reduce training cost. The results are shown in Table 4. The difference when using different variations of verbalization was small. However, verbalizing available features did show slightly higher performance (macro-F_1_ = 0.662), especially in terms of precision, while verbalizing missing features had less of a positive effect overall. In contrastive loss experiments, the model trained jointly using BCE and unsupervised contrastive loss from SimCSE^32^ had the best performance (0.668 macro-F_1_), while models using label-aware contrastive loss significantly underperformed this model.

**Table 4:**
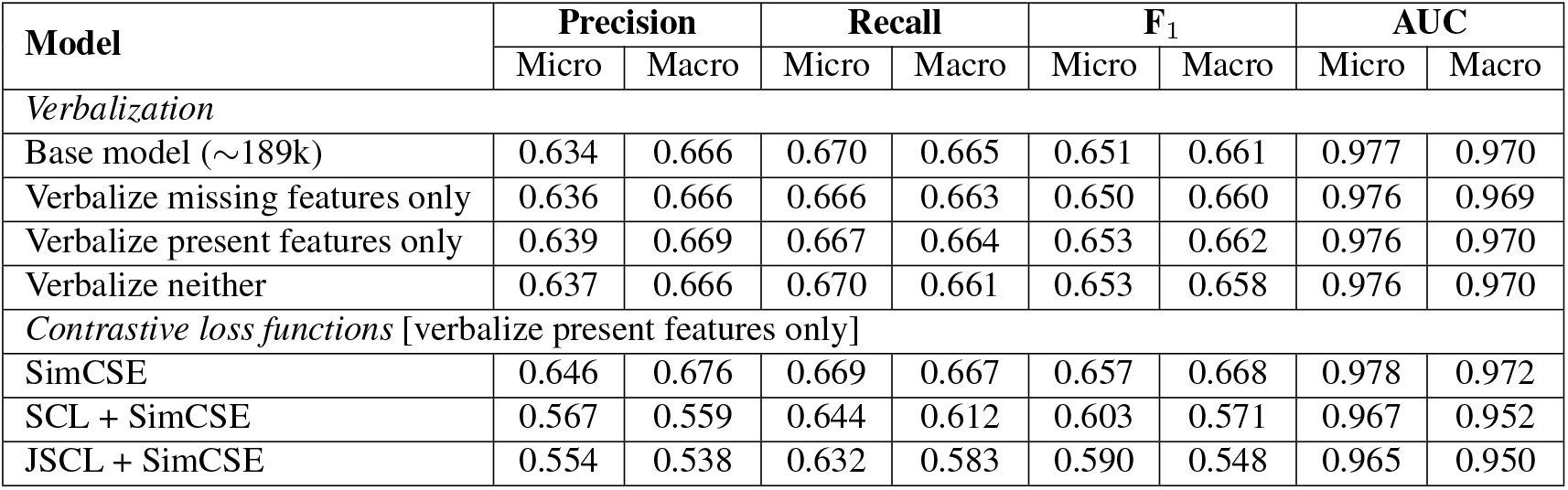
Ablation study for verbalizing features and different contrastive loss functions. The base model (189k) verbalizes both present and missing features. Contrastive loss experiments verbalize present features only.

| Model | Precision |  | Recall |  | F<sub>1</sub> |  | AUC |  |
| --- | --- | --- | --- | --- | --- | --- | --- | --- |
|  | Micro | Macro | Micro | Macro | Micro | Macro | Micro | Macro |
| *Verbalization* |  |  |  |  |  |  |  |  |
| Base model (~189k) | 0.634 | 0.666 | 0.670 | 0.665 | 0.651 | 0.661 | 0.977 | 0.970 |
| Verbalize missing features only | 0.636 | 0.666 | 0.666 | 0.663 | 0.650 | 0.660 | 0.976 | 0.969 |
| Verbalize present features only | 0.639 | 0.669 | 0.667 | 0.664 | 0.653 | 0.662 | 0.976 | 0.970 |
| Verbalize neither | 0.637 | 0.666 | 0.670 | 0.661 | 0.653 | 0.658 | 0.976 | 0.970 |
| *Contrastive loss functions* [verbalize present features only] |  |  |  |  |  |  |  |  |
| SimCSE | 0.646 | 0.676 | 0.669 | 0.667 | 0.657 | 0.668 | 0.978 | 0.972 |
| SCL + SimCSE | 0.567 | 0.559 | 0.644 | 0.612 | 0.603 | 0.571 | 0.967 | 0.952 |
| JSCL + SimCSE | 0.554 | 0.538 | 0.632 | 0.583 | 0.590 | 0.548 | 0.965 | 0.950 |

## Discussion

In this work, we aimed to improve on previous work (i.e., MultiTagger^19^) by using more modern machine learning methods and de-emphasizing extensive feature engineering. The PubMedBERT-based models performed relatively well overall; the best results were obtained with a model that used 80% undersampling and feature verbalization. This model outperformed MultiTagger, increasing macro-F_1_ from 0.416 to 0.632, as did a baseline PubMedBERT model trained on a smaller training set (macro-F_1_: 0.618), indicating the strength of BERT-based models in capturing contextual, PT-related information. Considering that MultiTagger was trained on human-related articles only, we also compared the performance on the subset of articles with the MeSH term Humans (n=50,656). We observed similar trends, although the difference was smaller (0.422 vs. our 0.589 macro-F_1_).

Undersampling was effective for training PubMedBERT models, which require significant computational resources especially with the large training sets that we constructed. Undersampling also led to a more balanced distribution of labels, which was beneficial for model training. Reducing the minimum number of instances per label more drastically to 150 had a negative effect, as there were fewer positive instances for all PTs and not just the majority classes.

The effect of using feature verbalization was mixed. Verbalizing missing features did not have much effect. While verbalizing available features had the highest performance overall, the differences were relatively small. Because verbalizing features is trivial, additional metadata features could be considered to improve performance further. In verbalizing features, we were inspired by MultiTagger, but we did not consider some features that required more complex processing (e.g., journal similarity feature^37^). Including these features could also be helpful in improving results. We have not assessed the contributions of individual metadata features, which we leave for future work.

Somewhat to our surprise, supervised contrastive learning degraded performance, while SimCSE (unsupervised) had a positive effect. Contrastive learning optimizes two properties of high-quality representations: uniformity and alignment^38^. Unsupervised SimCSE focuses on uniformity, which refers to how close the instances are in the embedding space^38^, and acts as a form of regularization through data augmentation^32^. On the other hand, our label-aware methods (SCL and JSCL) more directly impact alignment, which refers to how close positive pairs of instances are. Alignment in multi-label settings is far more difficult than in binary classification, which may have led to poor performance. Our use of a single static anchor pair may have affected the performance as well, because multiple instances may be needed to triangulate the optimal alignment space. Additionally, we used the same hyperparameters, temperature (0.05) and *α* (0.4) in all experiments. Further hyperparameter tuning should be considered in future work. Lastly, the unsupervised SimCSE method should be evaluated on different training set sizes. Due to time constraints, these experiments have been left to future work.

### Limitations

The PT labels we used are human-curated, but errors do exist in the labels. For example, many RCTs are not annotated with the Randomized Controlled Trial PT in MEDLINE^16,39^. Another limitation is that the PTs included in MEDLINE are not exhaustive; they do not represent all the different kinds of research methodologies and study designs found in medicine and life sciences. For example, the dataset has few labels dedicated to animal studies, e.g., pre-clinical research involving animal models of disease. There are also no specific labels focusing on molecular biology/chemistry research. In future work, we plan to address some of these issues by considering an expanded set of PTs. Additionally, the classification task may not be how these models are best utilized in real world settings. For information retrieval specifically, probabilistic scores, rather than binary labels, may be more useful for users (e.g., allowing adjustable thresholds during search). MultiTagger does this, while our models do not because the scores are not yet calibrated. To maximize utility, our scores should be calibrated in future work.

## Conclusions

In this study, we built on previous work^19^ to generate a large dataset of PubMed articles labeled across 59 PTs using a combination of PubMed queries and MEDLINE indexing terms. Based on this dataset, we trained and validated PubMedBERT-based document classification models, augmented with techniques like undersampling, feature verbalization, and contrastive learning. Our results show that these models significantly outperform MultiTagger. Future work will focus on expanding the dataset to include PTs not currently considered in MEDLINE (e.g., diagnostic accuracy test studies), incorporating full-text features as well as other metadata-related features, calibrating models, and improving model performance and interpretability.

## Data Availability

We make our data and code available at https://github.com/ScienceNLP-Lab/MultiTagger-v2/tree/main/AMIA.

## Acknowledgments

We thank Arthur Holt for providing MultiTagger predictions, as well as Shufan Ming and Shruthan Radhakrishna for reviewing the code used for model development and evaluation. This work was supported by the National Library of Medicine of the National Institutes of Health under the award number R01LM14292. The content is solely the responsibility of the authors and does not necessarily represent the official views of the National Institutes of Health. The funder had no role in considering the study design or in the collection, analysis, interpretation of data, writing of the report, or decision to submit the article for publication.

